## Supplementary appendix for "Rapid antigen testing as a reactive response to surges in nosocomial SARS-CoV-2 outbreak risk"

### I. Simulating SARS-CoV-2 outbreaks using CTCmodeler

SARS-CoV-2 outbreaks were simulated using CTCmodeler, a stochastic, individual-based transmission model coded in C++. This peer-reviewed[1–3] model simulates pathogen transmission along contact networks describing inter-individual interactions among patients and 13 types of staff (e.g. nurse, physiotherapist) in a 170-bed, five-ward LTCF setting. Model contacts are fit to real high-resolution close-proximity interaction data (n=2.67 million distinct person-person interactions). Interaction data were recorded by wearable sensors carried by a cumulative 329 patients and 261 members of staff over four months in 2009 in a rehabilitation hospital in northern France. These data have been described and analyzed in detail elsewhere.[4,5] Among other observations, healthcare workers were found to contact many distinct patients each day, spending more time with older and higher-needs patients. Patients, however, had particularly high rates of contact with other patients, reflecting the group meals, art classes and other social activities that took place over the data collection period in this pre-pandemic LTCF.

The computer program underlying CTCmodeler is described in detail elsewhere.[1] In simulations, there were on average 170 patients and 240 members of staff present in the LTCF each week. Patient and staff interactions occurred over 30-second intervals, with contact frequencies and durations varying across the types of interactions that take place (e.g. patient-patient, patient-nurse), in which of the facility's five wards the individuals are found (e.g. geriatrics, nutrition), and at what times of day and days of the week interactions occur. Synthetic contact data fit to these individual-level data were generated previously, stratified by hour of the day, day of the week, ward, and type of individual (see Smith *et al.*).[3] These synthetic data are used in the present work, hence no sensitive human data were used in this study.

We previously applied CTCmodeler to simulate SARS-CoV-2 transmission and evaluate efficacy and efficiency of RT-PCR testing strategies in the context of limited testing resources (see Smith *et al.*).[3] Three key modelling assumptions were made to reflect SARS-CoV-2 epidemiology, informed using epidemiological data and parameter estimates from the literature. First, SARS-CoV-2 was assumed to be introduced into the LTCF through either newly admitted patients or staff members infected in the community. Second, infection followed a modified Susceptible-Exposed-Infected-Recovered process, including *pre-symptomatic* and *asymptomatic* infection, with stochastic progression through infection stages. Third, transmission could occur during contacts between infectious and susceptible individuals, with transmission probability scaling linearly with contact duration, until saturating for contacts exceeding 60 minutes. An Overview, Design concepts, and Details (ODD) protocol for individual-based modelling describing this application of CTCmodeler to SARS-CoV-2 is provided in Smith *et al.*[3]

Here, a range of additional assumptions were integrated into simulations to reflect the evolving epidemiology of COVID-19 and present research context. These include:

(i) index cases acquired during a surge in community SARS-CoV-2 circulation,
(ii) heterogeneous SARS-CoV-2 introductions from the community reflecting local infection burden,
(iii) variable transmissibility according to symptom status,
(iv) immediate isolation of patients with severe COVID-19 symptoms,
(v) sick-leave and shift replacement for staff with severe COVID-19 symptoms,
(vi) a risk of healthcare workers acting as transient SARS-CoV-2 vectors,
(vii) initialization conditions reflecting an ongoing pandemic context,
(viii) implementation of public health interventions for SARS-CoV-2 transmission prevention ("COVID-19 containment measures").

These new assumptions are detailed below, with model parameters provided in Supplementary table S1.

1. *Index cases acquired during a surge in community SARS-CoV-2 circulation*

We distinguish between SARS-CoV-2 index cases and SARS-CoV-2 introductions. Index cases were defined as patients and staff infected with SARS-CoV-2 upon simulation outset, who were thus assumed to have acquired infection prior to the simulation period. These infections were conceptualized as resulting from inter-generational mixing in the community over festive holidays over the week prior to simulation outset, leading to an increase in nosocomial outbreak risk within the LTCF. Upon simulation initialization, we assume that 50% of patients (n=85) and 100% of staff (n=240) had contacts in the community over the previous week, with a 1.2% probability of acquiring SARS-CoV-2. This translated to 1 index patient and 3 index members of staff infected with SARS-CoV-2 at simulation outset, randomly selected among all patients and staff present in the LTCF. Index cases were assumed to be in any infection stage except *severe symptomatic*. Calibrated using model transition parameters (Supplementary table S1), we assumed that 28.0% of index cases were *exposed*, 4.8% *pre-asymptomatic*, 16.8% *asymptomatic*, 11.2% *pre-symptomatic* and 39.2% *mild symptomatic*.

2. *Heterogeneous SARS-CoV-2 introductions from the community reflecting local infection burden*

Introductions were defined as subsequent cases of SARS-CoV-2 infection introduced into the LTCF from the community over the course of simulation time, limited to staff members (assumed to contact individuals in the community outside work hours) and newly admitted patients (assumed to potentially carry the virus upon LTCF entry).

We assumed that staff introductions were new infections (in the *exposed* stage) acquired in the previous 24 hours. This reflects infection resulting from community contacts since that individual's previous shift. Community SARS-CoV-2 infection incidence was used to calculate the daily probability of any working member of staff becoming infected with the virus and introducing it to the LTCF. Using French epidemic data from late January 2021 (daily incidence of 26,676 cases among a population of 67.1 million individuals), we estimated a daily incidence rate of 0.04%. In a "high incidence" sensitivity analysis, we assumed under-reporting of 90.8% (as estimated elsewhere by Anand *et al.*), translating to an incidence rate of 0.37%.[6]

For patient introductions we assumed that infection is acquired at any time prior to LTCF admission. We used community SARS-CoV-2 prevalence to estimate the probability that a new patient entering the LTCF is already infected. Using the incidence data above and assuming an infection duration of 12.5 days (the mean duration under our modelling assumptions), we estimated a community SARS-

CoV-2 infection prevalence of 0.50%, or 4.57% in the “high incidence” sensitivity analysis. Patient introductions could be in any infection stage, using the same stage-specific probabilities as for index cases (above).

Combining patient and staff introductions, there were a mean 0.08 introductions/day in the baseline “low incidence” scenario, and a mean 0.8 introductions/day in a “high incidence” sensitivity analysis.

3. *Variable transmissibility according to symptom status*

Previously, we estimated a SARS-CoV-2 transmission rate per minute of infectious contact $p$=0.14% (see theoretical discussion in Temime *et al*. and its application in Smith *et al*.).[3,7] Under our assumptions, the probability of SARS-CoV-2 transmission from an infectious individual $i$ to a susceptible individual $j$ ($P_{i\to j}$) depends on the duration of their contact in minutes ($D_{i,j}$), given by

$$P_{i\to j} = p \times D_{i,j} \quad (1)$$

Here, we now assume that SARS-CoV-2 infectivity varies between *symptomatic* (including *pre-symptomatic*) and *asymptomatic* (including *pre-asymptomatic*) infections. In a systematic review and meta-analysis, Buitrago-Garcia *et al*. estimated the secondary attack rate among contacts of asymptomatic infections to be 35% relative to symptomatic infections, and that approximately 30% of all infections remained asymptomatic.[8] Using these data, we stratified $p$ to estimate distinct transmission rates from *symptomatic* ($p_{sym}$) and *asymptomatic* ($p_{asym}$) individuals as

$$0.7 \times p_{sym} + 0.3 \times p_{asym} = 0.14\% \quad (2)$$

where

$$p_{asym} = 0.35 \times p_{sym} \quad (3)$$

Solving these equations, $p_{sym}$ = 0.174% and $p_{asym}$ = 0.061%.

4. *Patients with severe COVID-19 symptoms: isolation*

We assumed that patients with severe COVID-19 symptoms are automatically isolated (independent of retrospective surveillance and isolation interventions introduced and evaluated later). We assumed a lag from symptom onset to isolation (24 hours), 100% isolation efficacy for transmission prevention ($p_{sym,isolated}$ = 0, $p_{asym,isolated}$ = 0), and an isolation duration equivalent to the remaining duration of infection (i.e. duration of symptoms), drawn from log-normal(7,7) distribution.

5. *Staff with severe COVID-19 symptoms: sick-leave and shift replacement*

We assumed that staff with severe COVID-19 symptoms immediately go on sick leave, with a duration equivalent to symptom duration (drawn from log-normal(7,7) distribution), after which they return to the LTCF *recovered*. During sick-leave, staff were replaced with a temporary member of staff who executes the same functions within the LTCF, and hence with no change to the underlying contact network. Probability of SARS-CoV-2 infection among newly arrived temporary replacement staff is calculated using community prevalence data (as for patient introductions, see above).

*6. Healthcare workers as transient SARS-CoV-2 vectors*

We allowed members of staff to act as transient vectors for SARS-CoV-2 after contact with infectious patients.[9] Transient vectors were assumed to physically "carry" SARS-CoV-2, and could transmit the virus to subsequent patients visited in quick succession (within 60 minutes), infecting those patients without themselves becoming infected. The probability of a member of staff *i* becoming a vector ($P_v$) was assumed to depend on the duration of their contact with the infectious patient *j*, $D_{i,j}$. This is given by

$$P_v = p_{carriage} \times D_{i,j} \times (1 - P_{protection}) \quad (4)$$

and the probability of this transient vector *i* then infecting a susceptible patient *k* ($P_{i \to k}$) during a subsequent contact relies on transmission rate *p* (using *p* to represent either $p_{sym}$ or $p_{asym}$), thus taking the same form as standard host-to-host transmission, given by

$$P_{i \to k} = p \times D_{i,k} \quad (5)$$

but limited to 60 minutes from the end of contact with *j*, after which time *i* loses transient carriage and is no longer a vector.

Here, $p_{carriage}$ is the per-minute probability of a member of staff acquiring transient SARS-CoV-2 carriage. We set a saturation threshold at 80% such that transient carriage is not inevitable subsequent to long infectious contacts. Similar to transmission rates $p_{sym}$ and $p_{asym}$ (see above), we assumed that $p_{carriage}$ varies between *symptomatic* and *asymptomatic* patients, such that $p_{carriage\_asym} = 0.35 \times p_{carriage\_sym}$. Finally, $P_{protection}$ describes the degree to which staff protect themselves during interactions with patients. The latter reflects an assumed asymmetry in patient-staff interactions, in which staff take measures to protect themselves from perceived risk of acquiring the virus when caring for patients, but take less care to protect patients from virus potentially lingering on their clothing or equipment.

*7. Initialization conditions for pandemic context*

In addition to the index cases introduced above, we updated two key initialization conditions for outbreak simulation to reflect the ongoing pandemic context of COVID-19. First, we assumed a baseline immunizing seroprevalence of 20% among patients and staff. For each individual in each outbreak simulation, initial infection status was determined stochastically, with a 20% probability of entering the simulation already *recovered*. Second, we assumed that baseline rates of infection prevention and control were improved relative to pre-pandemic baseline. This reflects observed increases in compliance to hygiene and infection prevention measures in healthcare institutions worldwide since the beginning of the COVID-19 pandemic.[10–12] For this, we introduced the parameter $P_{IPC}$, which modifies rates of transmission ($P_{i \to j,IPC}$) and transient carriage acquisition ($P_{v,IPC}$) across all patients and staff, given by:

$$P_{i \to j,IPC} = P_{i \to j} \times (1 - P_{IPC}) \quad (6)$$

and

$$P_{v,IPC} = P_v \times (1 - P_{IPC}) \quad (7)$$

Using data from a clinical trial across 33 Dutch nursing homes, we assumed a baseline 36% compliance to IPC measures in such an intervention context ($P_{IPC}$ = 0.36).[13]

8. *COVID-19 containment measures: public health interventions for SARS-CoV-2 transmission prevention*

We included three different COVID-19 containment measures in outbreak simulations.

First, we included a patient social distancing intervention, interpreted as cancellation of all social activities occurring in the baseline pre-pandemic contact network. This was modelled by removing all contacts involving ≥3 patients simultaneously. Impacts of this intervention on dynamic contact behaviours simulated by CTCmodeler are visualized in Supplementary figure S1. We did not impose staff social distancing, under the assumption that staff contacts are necessary for provisioning of care.

Second, we considered a mandatory face mask policy among all patients and staff. In a systematic review and meta-analysis, Liang *et al.* estimated an 80% reduction in risk of respiratory virus transmission through wearing of face masks.[14] This was modelled here by setting $P_{IPC}$ = 0.8, reducing rates of transmission and transient carriage acquisition across patients and staff.

Third, we included a partial vaccination intervention, in which we assumed a 50% rate of immunizing seroprevalence (relative to 20% baseline). This could be interpreted in different ways, for instance as 100% vaccine coverage for a vaccine providing 50% protection from infection with the locally circulating strain (in the absence of any naturally-acquired immunity), 75% vaccine coverage for a vaccine providing 75% protection from infection, etc.

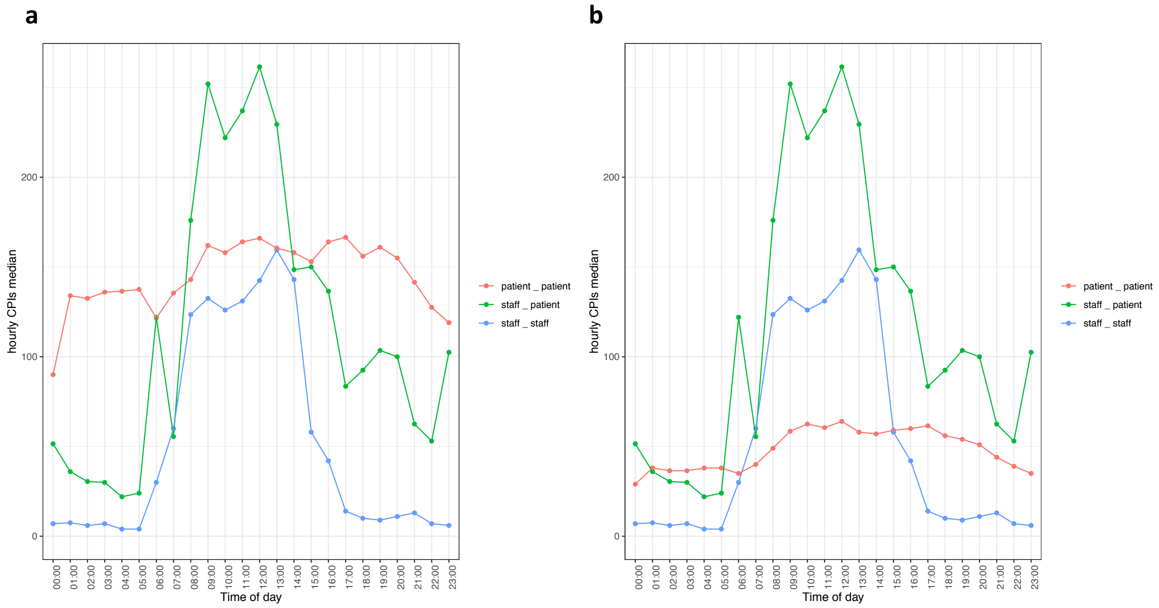


**Supplementary figure S1.** Impacts of the patient social distancing intervention on daily contact behaviour. Over the course of one day (x-axis), the median hourly number of close-proximity interactions (CPIs, y-axis) is shown. Patient-patient contacts vary considerably across **(a)** the baseline LTCF and **(b)** the LTCF with the patient social distancing intervention in place, while patient-staff and staff-staff contacts are unchanged.

**Supplementary table S1.** Transmission model parameter estimates. COVID-19 infection parameters are unchanged since previous publication in Smith *et al.*[3]

| Parameter | Value [distribution] | Source |
|---|---|---|
| **COVID-19 infection parameters** | | |
| Duration of exposed period (latency) | 2-5 days [uniform] | Approximated from Lauer et al. 2020 and Wei et al. 2020 [15,16] |
| Duration of pre-symptomatic or pre-asymptomatic period | 1-3 days [uniform] | Approximated from Lauer et al. 2020 and Wei et al. 2020 [15,16] |
| Duration of symptomatic period (whether asymptomatic, mild symptomatic or severe symptomatic) | 7 days [log-normal, $\sigma^2$ = 7] | Approximated from He et al. 2020 [17] |
| Proportion of COVID-19 infections presenting any symptoms | 0.7 | Buitrago-Garcia et al. 2020 [8] |
| Proportion of symptomatic COVID-19 infections with severe symptoms | 0.2 | Wu et al. 2020 [18] |
| Daily incidence proportion of non-COVID but COVID-like symptoms | 0.011 | Estimated from OSCOUR data, described in Fouillet et al. 2015 [19] |
| **SARS-CoV-2 transmission parameters** | | |
| SARS-CoV-2 transmission rate per minute of contact, pre-symptomatic and symptomatic infection ($p_{sym}$) | 0.001739 | Previous estimate in Smith et al. 2020 scaled using data from Buitrago-Garcia et al. 2020 [3,8] |
| SARS-CoV-2 transmission rate per minute of contact, pre-asymptomatic and asymptomatic infection ($p_{asym}$) | 0.000609 | Previous estimate in Smith et al. 2020 scaled using data from Buitrago-Garcia et al. 2020 [3,8] |
| Staff SARS-CoV-2 transient carriage acquisition rate per minute of contact with symptomatic or pre-symptomatic patients ($p_{carriage\_sym}$) | 0.1 | Assumed |
| Staff SARS-CoV-2 transient carriage acquisition rate per minute of contact with asymptomatic or pre-asymptomatic patients ($p_{carriage\_asym}$) | 0.035 | Assumed |
| Degree of patient and staff compliance to infection prevention and control interventions ($P_{IPC}$) | 0.36 (0.80 under face mask intervention) | Teesing et al. 2020 (Liang et al. 2020) [13,14] |
| Degree to which staff protect themselves (but not patients) from high-risk contacts potentially leading to SARS-CoV-2 transmission ($P_{protection}$) | 0.80 | Assumed |
| **SARS-CoV-2 introductions from the community** | | |
| Daily probability of SARS-CoV-2 introduction from staff | 0.03976% (0.366% in sensitivity analysis) | Calibrated to incidence in France in January 2021 [20] |
| Probability of SARS-CoV-2 introduction per new patient admission | 0.497% (4.57% in sensitivity analysis) | Calibrated to incidence in France in January 2021 [20] |
| Probability of SARS-CoV-2 introduction per replacement staff | 0.497% (4.57% in sensitivity analysis) | Calibrated to incidence in France in January 2021 [20] |
| Probability exposed (patient admissions, replacement staff) | 0.28 | From infection parameters |
| Probability pre-asymptomatic (patient admissions, replacement staff) | 0.048 | From infection parameters |
| Probability asymptomatic (patient admissions, replacement staff) | 0.168 | From infection parameters |
| Probability pre-symptomatic (patient admissions, replacement staff) | 0.112 | From infection parameters |
| Probability symptomatic mild (patient admissions, replacement staff) | 0.392 | From infection parameters |

### II. Supplementary results: SARS-CoV-2 outbreak simulations

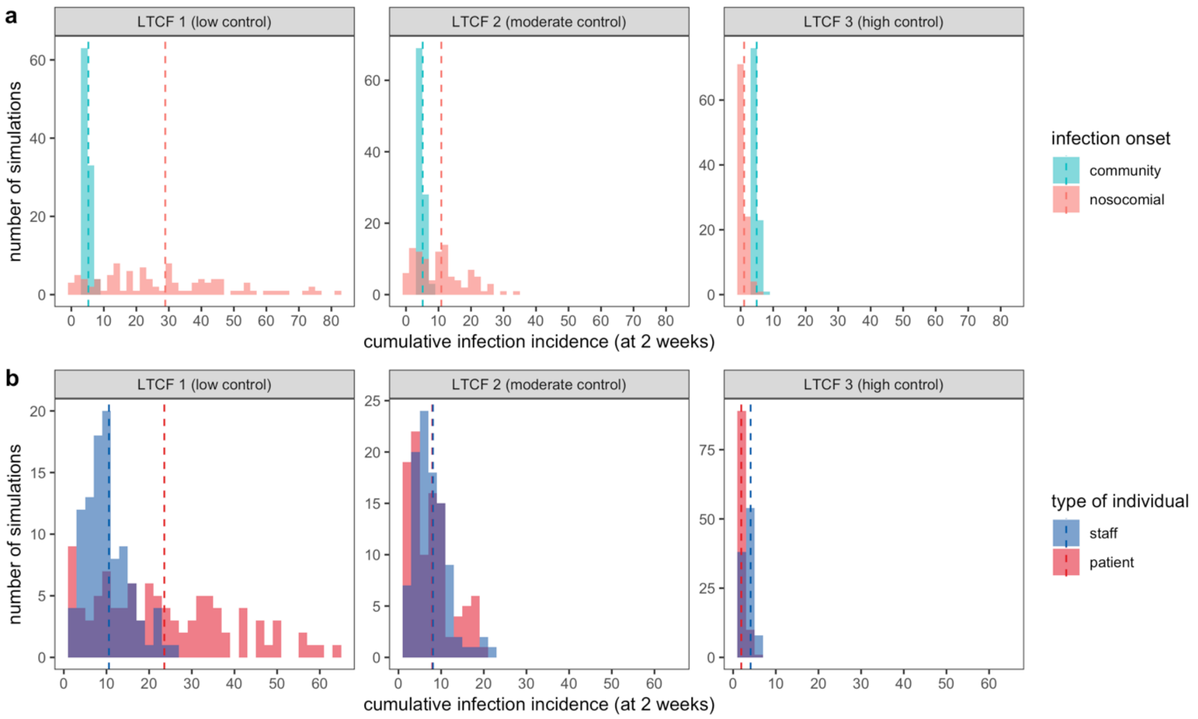


**Supplementary figure S2.** Across LTCFs (columns), the distributions of cumulative SARS-CoV-2 infection incidence at two weeks, stratified by **(a)** location of infection onset, and **(b)** type of individual infected. Dashed vertical lines represent means across 100 outbreak simulations. LTCF = long-term care facility.

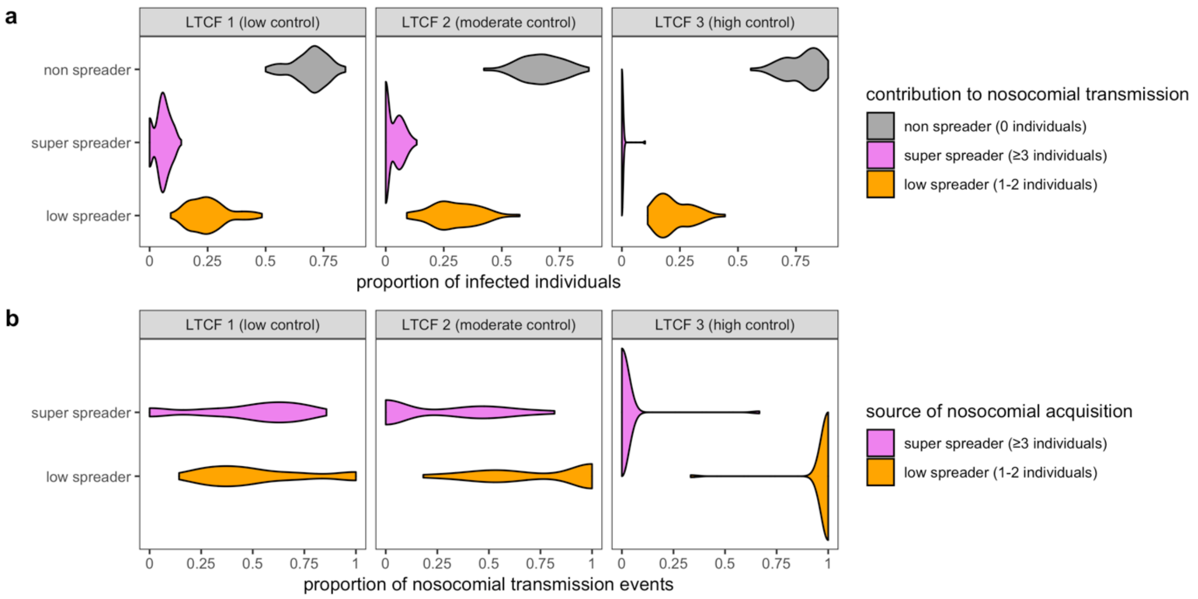


**Supplementary figure S3.** Impact of super-spreading on nosocomial SARS-CoV-2 outbreaks. **(a)** Contribution of super-spreading to nosocomial transmission: across simulated outbreaks, the distribution of the proportions of individuals that were super-spreaders (transmitted to ≥3 individuals, pink), low-spreaders (1-2 individuals, orange), and non-spreaders (0 individuals, grey). The proportion of infected individuals who never transmitted was higher in LTCF 3 (mean 77.7%) than in LTCFs 1 (69.4%) or 2 (67.0%). **(b)** Contribution of super-spreading to nosocomial acquisition: across outbreaks, the distribution of the proportions of acquisitions that resulted from super-spreaders (pink) versus low-spreaders (orange). LTCF = long-term care facility.

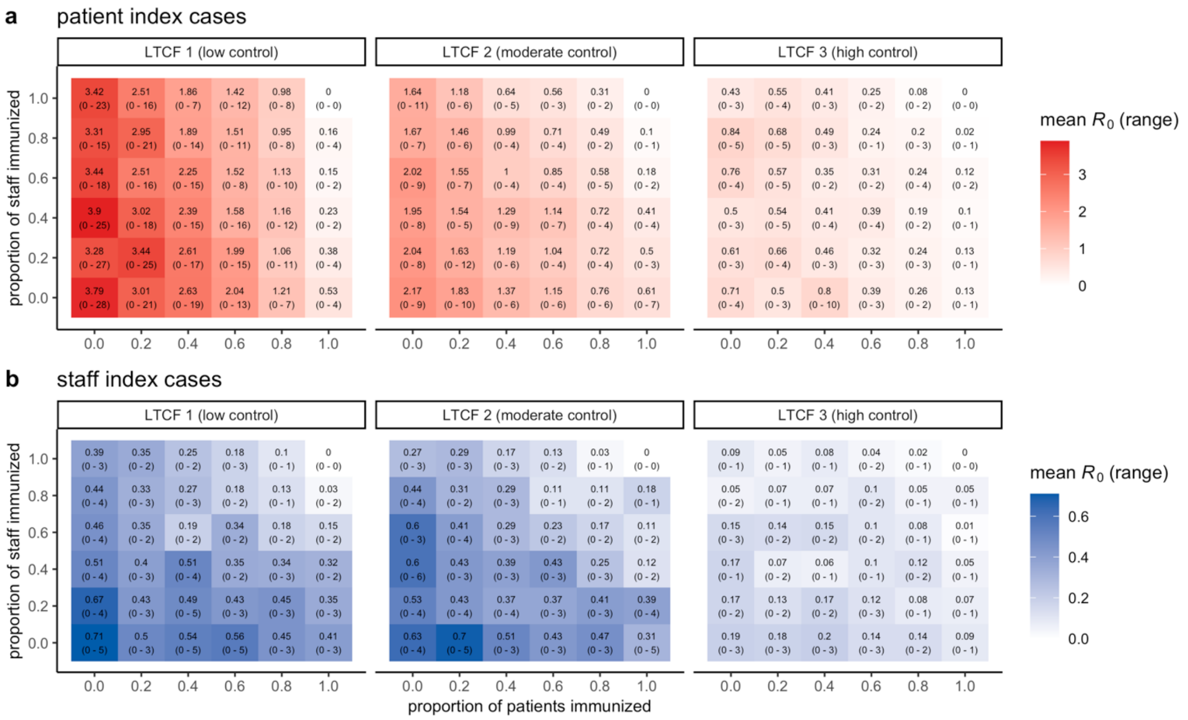

**Supplementary figure S4.** The mean (range) of the number of secondary nosocomial infections caused by index cases (for simplicity, $R_0$), stratified by **(a)** patient index cases and **(b)** staff index cases. Control measures for each LTCF are the same as presented in Figure 1, except for vaccination: here, the proportion of patients immunized at simulation outset is varied along the x-axis, and the proportion of staff along the y-axis. LTCF = long-term care facility.

### III. Simulating surveillance interventions using counterfactual analysis

*Surveillance interventions*

Final outputs from CTCmodeler simulations included a list of SARS-CoV-2 introductions from the community, a list of nosocomial transmission events (including donor and recipient IDs and mode of transmission, i.e. from an infected individual or transient vector), and the daily infection status of each unique individual in the LTCF. This information was used to inform a surveillance algorithm, which over simulation time identified which individuals were (i) newly admitted from the community, (ii) infected with SARS-CoV-2, and (iii) experiencing COVID-19 symptoms. Using these data, tests were allocated according to the different surveillance interventions listed in Supplementary table S2. To account for intervention heterogeneity, we varied screening targets (*patients & staff*, only patients, only staff), the type of test used for screening (*Ag-RDT A*, Ag-RDT B, RT-PCR), and the shape of diagnostic sensitivity curves (*time-varying*, uniform, perfect) (italicized items represent baseline assumptions used in the main analysis, unless stated otherwise).

*Counterfactual scenarios*

Surveillance interventions were applied retrospectively to daily outbreak data for precise estimation of intervention effects, using methods adapted from single-world counterfactual analysis (see Kaminsky *et al*.).[21] Each stochastic "run" of the surveillance algorithm was conducted in five steps. First, the algorithm was applied to each outbreak to determine who to test for SARS-CoV-2 infection and when, and using which type of test, according to the surveillance interventions described in Supplementary table S2. Second, test results were determined stochastically, with probability of SARS-CoV-2 detection depending on test sensitivity $s(t)$ (sensitivity functions are introduced below). Third, individuals were retrospectively "isolated" upon SARS-CoV-2 diagnosis (positive test result), assuming immediate isolation for Ag-RDT but a 24-hour lag for RT-PCR (reflecting a lag between sample and result). Fourth, *counterfactual scenarios* were simulated by pruning transmission events occurring subsequent to isolation, i.e. removing all transmission chains originating from isolated individuals. Fifth, nosocomial incidence was re-calculated subsequent to transmission pruning, and surveillance outcomes were calculated as described in the main text. This process was repeated n=100 times for each outbreak.

With these methods, in each run of surveillance "uncontrolled" epidemics with no surveillance were matched to "controlled" counterfactual scenarios with surveillance, facilitating single-world comparison of intervention efficacy across stochastic outbreak simulations (illustrated in Figure 3). This methodology was particularly important for interventions #11 to #27, which introduced screening against a backdrop of a facility already conducting routine testing. This resulted in counterfactual scenarios with multiple "levels" of surveillance, in which the efficacy and efficiency of screening is interpreted as relative to counterfactual scenarios with routine testing already in place (and not relative to baseline uncontrolled epidemics). For these calculations, routine testing was always simulated first, with downstream transmission chains pruned as described above; second, screening was applied, with potential only to prune remaining transmission chains not already pruned by routine RT-PCR; and third, if conducting a second round of screening, the only chains potentially pruned were those not already pruned by routine testing and the first round of screening. In this way, infections averted by subsequent levels of surveillance did not double-count infections already averted by previous levels.

**Supplementary table S2.** List of surveillance interventions considered. Italicized text indicates distinct stratification of columns across routine testing and screening interventions, and alternative stratifications in sensitivity analysis.

| # | Surveillance category | Screening timing | Test used | Test target |
|---|---|---|---|---|
| **1** | Routine testing | / | RT-PCR | Individuals with COVID-like symptoms and new patient admissions |
| **2** | 1-round screening | Day 1 | Ag-RDT (A)<br><br>*In sensitivity analysis:*<br>Ag-RDT (B), RT-PCR | Patients & staff<br><br>*In sensitivity analysis:*<br>only patients, only staff |
| **3** | | Day 2 | | |
| **4** | | Day 3 | | |
| **5** | | Day 4 | | |
| **6** | | Day 5 | | |
| **7** | | Day 6 | | |
| **8** | | Day 7 | | |
| **9** | | Day 8 | | |
| **10** | | Day 9 | | |
| **11** | Routine testing + 1-round screening | Day 1 | *For routine testing:*<br>RT-PCR<br><br>*For screening:*<br>Ag-RDT (A)<br><br>*For screening in sensitivity analysis:*<br>Ag-RDT (B), RT-PCR | *For routine testing:*<br>individuals with COVID-like symptoms and new patient admissions<br><br>*For screening:*<br>patients & staff<br><br>*For screening in sensitivity analysis:*<br>only patients, only staff |
| **12** | | Day 2 | | |
| **13** | | Day 3 | | |
| **14** | | Day 4 | | |
| **15** | | Day 5 | | |
| **16** | | Day 6 | | |
| **17** | | Day 7 | | |
| **18** | | Day 8 | | |
| **19** | | Day 9 | | |
| **20** | Routine testing + 2-round screening | Days 1 & 2 | *For routine testing:*<br>RT-PCR<br><br>*For screening:*<br>Ag-RDT (A)<br><br>*For screening in sensitivity analysis:*<br>Ag-RDT (B), RT-PCR | *For routine testing:*<br>individuals with COVID-like symptoms and new patient admissions<br><br>*For screening:*<br>patients & staff<br><br>*For screening in sensitivity analysis:*<br>only patients, only staff |
| **21** | | Days 1 & 3 | | |
| **22** | | Days 1 & 4 | | |
| **23** | | Days 1 & 5 | | |
| **24** | | Days 1 & 6 | | |
| **25** | | Days 1 & 7 | | |
| **26** | | Days 1 & 8 | | |
| **27** | | Days 1 & 9 | | |

*Diagnostic sensitivity of RT-PCR and Ag-RDT*

Test sensitivity, the probability of positive diagnosis for a true infection, was assumed to depend on infection age $t$, i.e. time since SARS-CoV-2 exposure. For nosocomial infections, infection age was calculated directly from transmission chains (model outputs). For community-onset infections, infection age upon LTCF introduction was generated stochastically depending on the stage of infection: $t = 1$ if exposed, $t \sim U(2,5) + 1$ if pre-symptomatic infectious, and $t \sim U(2,5) + U(1,3) + 1$ if symptomatic or asymptomatic infectious.

Test sensitivity as a function of infection age is expressed as $s_{PCR}(t)$ for RT-PCR and $s_{RDT}(t)$ for Ag-RDT. For RT-PCR, sensitivity estimates for $1 \leq t \leq 20$ were taken directly from a meta-analysis by Kucirka *et al.*,[22] with maximum sensitivity $s_{PCR} = 80.9\%$ at $t = 8$ days from SARS-CoV-2 exposure. We extrapolated beyond $t = 20$ days using a negative exponential function fit to days 11-20 (pink line in Supplementary figure S5). Consistent with findings from the literature, we assumed no difference in diagnostic sensitivity for symptomatic (including pre-symptomatic) and asymptomatic (including pre-asymptomatic) infections.[23–25]

Literature estimates for Ag-RDT sensitivity ($\alpha$) are typically expressed as relative to RT-PCR sensitivity, given here by $s_{RDT}(t) = \alpha \times s_{PCR}(t)$, such that absolute Ag-RDT sensitivity $s_{RDT}(t)$ varies in time with $s_{PCR}(t)$. In a meta-analysis of 73 clinical data sets and 31,202 samples, Brümmer *et al.* estimated Ag-RDT to be $\alpha = 73.8\%$ as sensitive as RT-PCR,[26] which when crossed with the data from Kucirka *et al*. corresponds to a peak absolute Ag-RDT sensitivity of $s_{RDT} = 59.7\%$ at $t = 8$ days. They found no statistical difference in Ag-RDT sensitivity between symptomatic and asymptomatic patients. However, they estimated greater relative Ag-RDT sensitivity up to one week from symptom onset ($\alpha = 87.5\%$) and lower relative sensitivity thereafter ($\alpha = 64.1\%$). We adjusted relative Ag-RDT sensitivity accordingly using two shape parameters $\gamma$ and $\beta$, and depending on time since peak RT-PCR sensitivity ($\tau = t - 8$), such that:

$$\alpha(t, \beta, \gamma) = (1 - \gamma) \times e^{-\beta \times \tau^2} \tag{8}$$

where sensitivity of Ag-RDT relative to RT-PCR varies with time, decreasing exponentially with increasing τ. Values of $\gamma$ and $\beta$ were estimated by minimizing the sum of three squared distance functions

$$\arg\min((d_1)^2 + (d_2)^2 + (d_3)^2) \tag{9}$$

using the R function optim. Assuming a 5-day incubation period, these correspond to distances of the target sensitivity function to estimates from Brümmer *et al*. for, respectively, sensitivity over all $t$ ($d_1$), up to $t \leq 11$ days from SARS-CoV-2 exposure ($d_2$), and $t > 11$ days from SARS-CoV-2 exposure ($d_3$). These are given by:

$$d_1 = 0.738 - \sum_{t=1}^{t_{max}} \frac{\alpha(t, \beta, \gamma)}{s_{RDT}(t)} \tag{10}$$

$$d_2 = 0.875 - \sum_{t=1}^{11} \frac{\alpha(t, \beta, \gamma)}{s_{RDT}(t)} \tag{11}$$

$$d_3 = 0.641 - \sum_{t=12}^{t_{max}} \frac{\alpha(t, \beta, \gamma)}{s_{RDT}(t)} \tag{12}$$

Solving $\alpha(t, \beta, \gamma)$ over all $t$ using the shape parameters estimated from the minimized sum of squared distances ($\beta = 0.001998$, $\gamma = 0.1172$) reproduced summary estimates from Brümmer *et al.* to within 0.3%: $\alpha_{t\leq 11} = 87.5\%$, $\alpha_{t>11} = 64.1\%$, and $\alpha_{t\geq 1} = 73.5\%$. This function was used to determine the probability of a positive test result for Ag-RDT testing as conducted in the main analysis (black line in Supplementary figure S5).

Alternative assumptions for RT-PCR and Ag-RDT diagnostic sensitivity considered in sensitivity analyses were: (i) uniform Ag-RDT sensitivity relative to time-varying RT-PCR ($\alpha = 73.8\%$, green line in Supplementary figure S5), (ii) uniform absolute sensitivity regardless of time since exposure for both RT-PCR ($s_{PCR} = 70\%$) and Ag-RDT ($s_{RDT} = 54\%$), and (iii) perfect absolute sensitivity for both ($s_{PCR} = s_{RDT} = 100\%$).

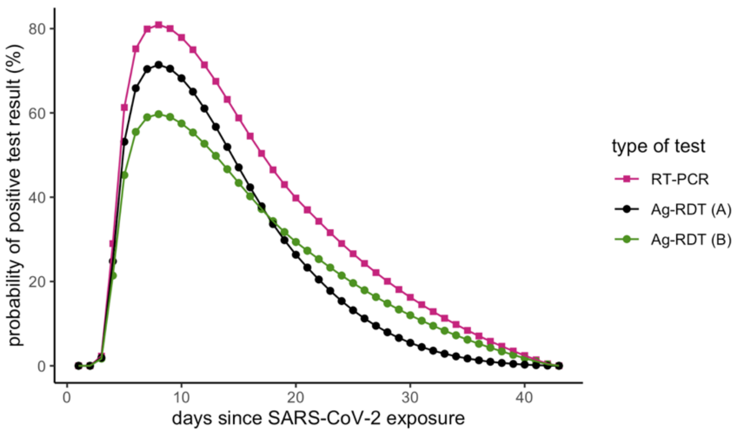


**Supplementary figure S5.** Test sensitivity as a function of time since infection, estimated by crossing data from meta-analyses by Kucirka *et al*. for RT-PCR (pink) and Brümmer *et al*. for Ag-RDT, considering time-varying Ag-RDT sensitivity relative to RT-PCR (black, for baseline analysis) and uniform relative sensitivity (green, for sensitivity analysis). RT-PCR = reverse transcriptase polymerase chain reaction; Ag-RDT = antigen rapid diagnostic testing.

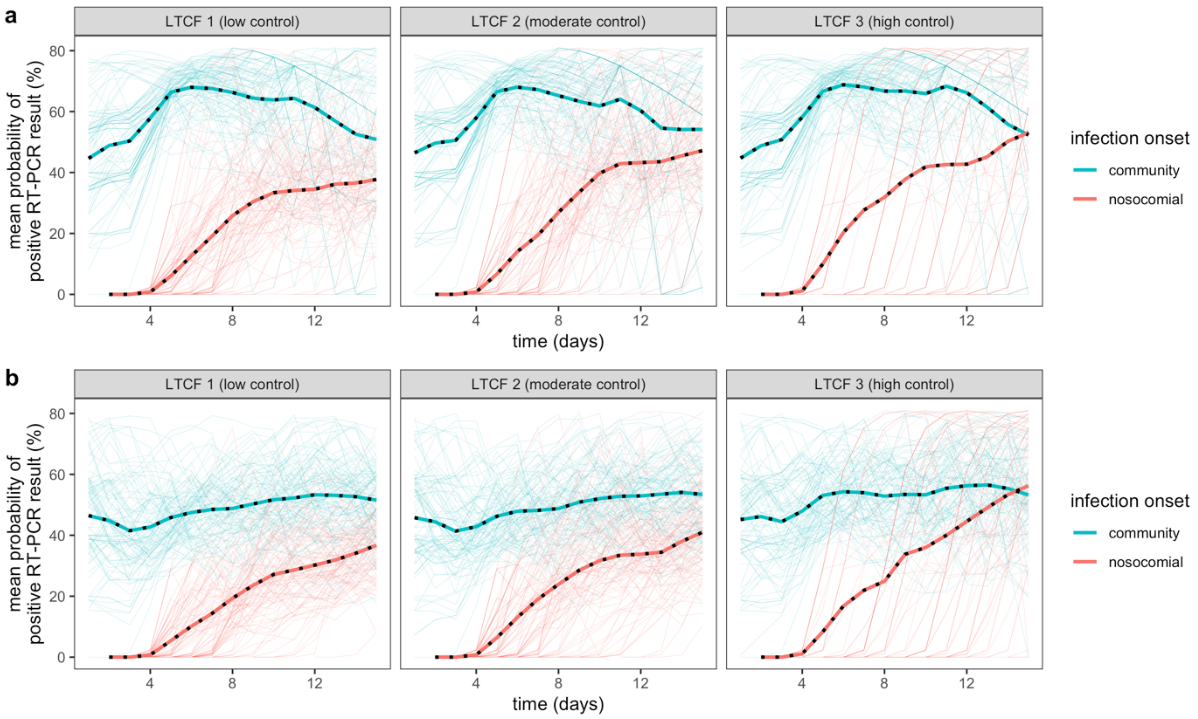


**Supplementary figure S6**. Temporal dynamics of RT-PCR test sensitivity among all individuals infected with SARS-CoV-2, in a hypothetical scenario of testing every infected individual every day. Test sensitivity is stratified by individuals who acquired infection in the community (blue) versus within the LTCF (red). Thin lines represent means for each outbreak simulation; thick lines represent means across all simulations. Sensitivity dynamics are shown for **(a)** the baseline low community incidence scenario, and **(b)** the high community incidence scenario. RT-PCR = reverse transcriptase polymerase chain reaction; LTCF = long-term care facility.

### IV. Supplementary results: surveillance efficacy and efficiency

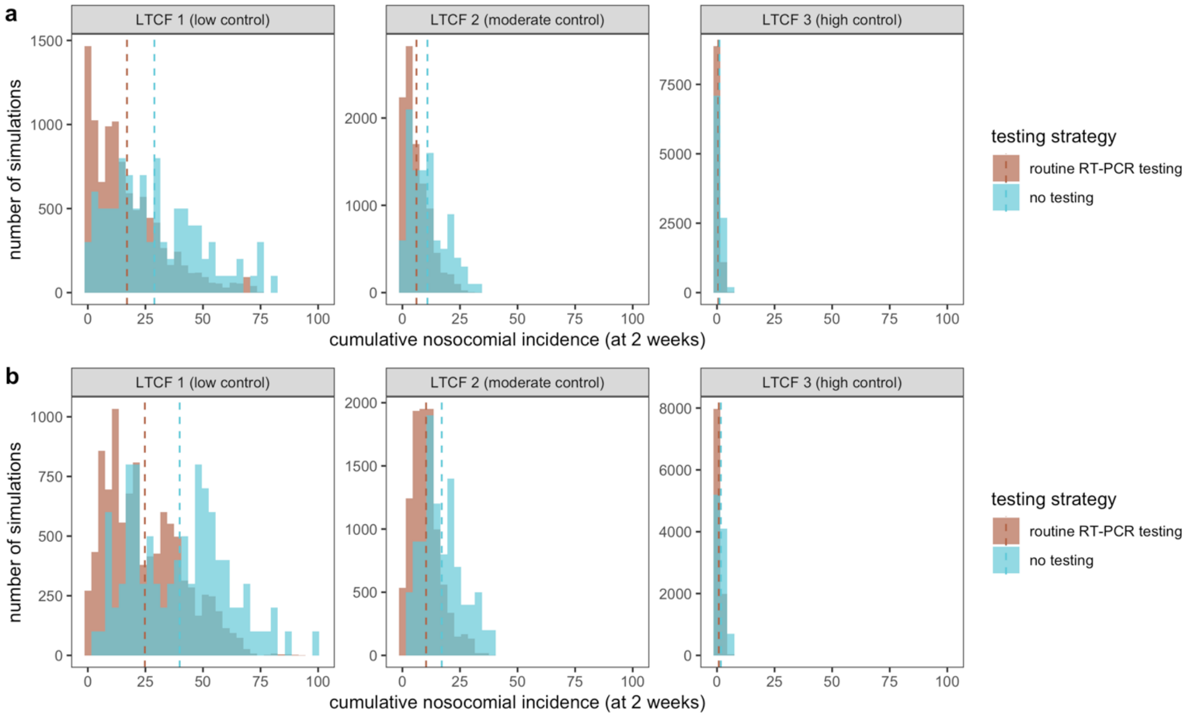


**Supplementary figure S7.** Compared to a scenario with no surveillance (blue), a counterfactual scenario with routine RT-PCR testing (brown) led to reduced cumulative nosocomial incidence (x-axis, with means as vertical dashed lines) due to pruning of transmission chains. For each LTCF (panels), relative efficacy (% reduction in incidence) was similar whether in **(a)** the baseline scenario of low community SARS-CoV-2 incidence, with a mean 1.1 new community-onset infections over two weeks subsequent to the initial surge; and **(b)** the high community incidence scenario, with a mean 10.6 new community-onset infections over two weeks. RT-PCR = reverse transcriptase polymerase chain reaction; LTCF = long-term care facility.

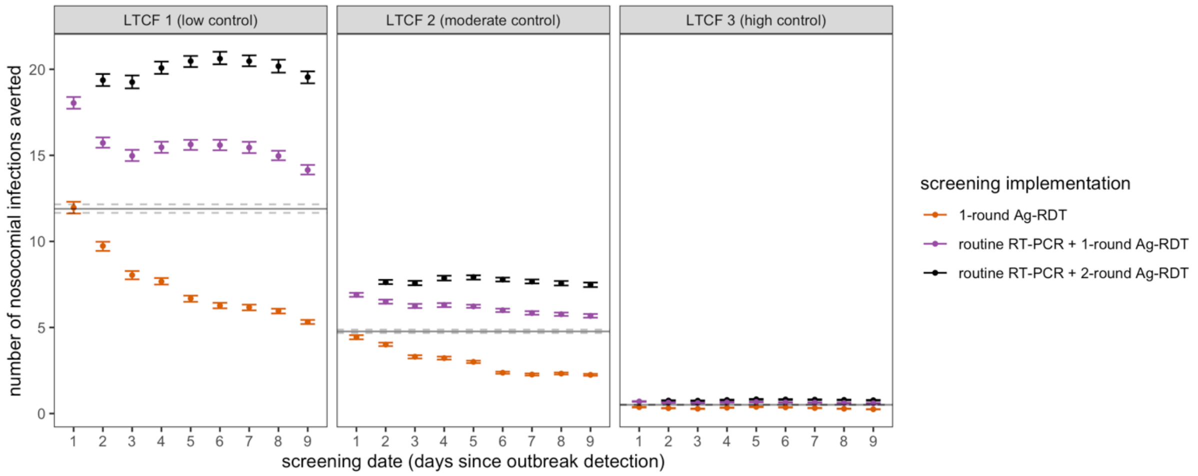


**Supplementary figure S8.** The total number of nosocomial SARS-CoV-2 infections averted by testing and screening interventions. This contrasts to relative reductions in incidence, as in Figure 2 in the main text. Points and error bars (solid horizontal lines and dashed lines, respectively, for routine RT-PCR testing) correspond to means and 95% confidence intervals estimated by bootstrap resampling (n=10,000). RT-PCR = reverse transcriptase polymerase chain reaction; Ag-RDT = antigen rapid diagnostic testing; LTCF = long-term care facility.

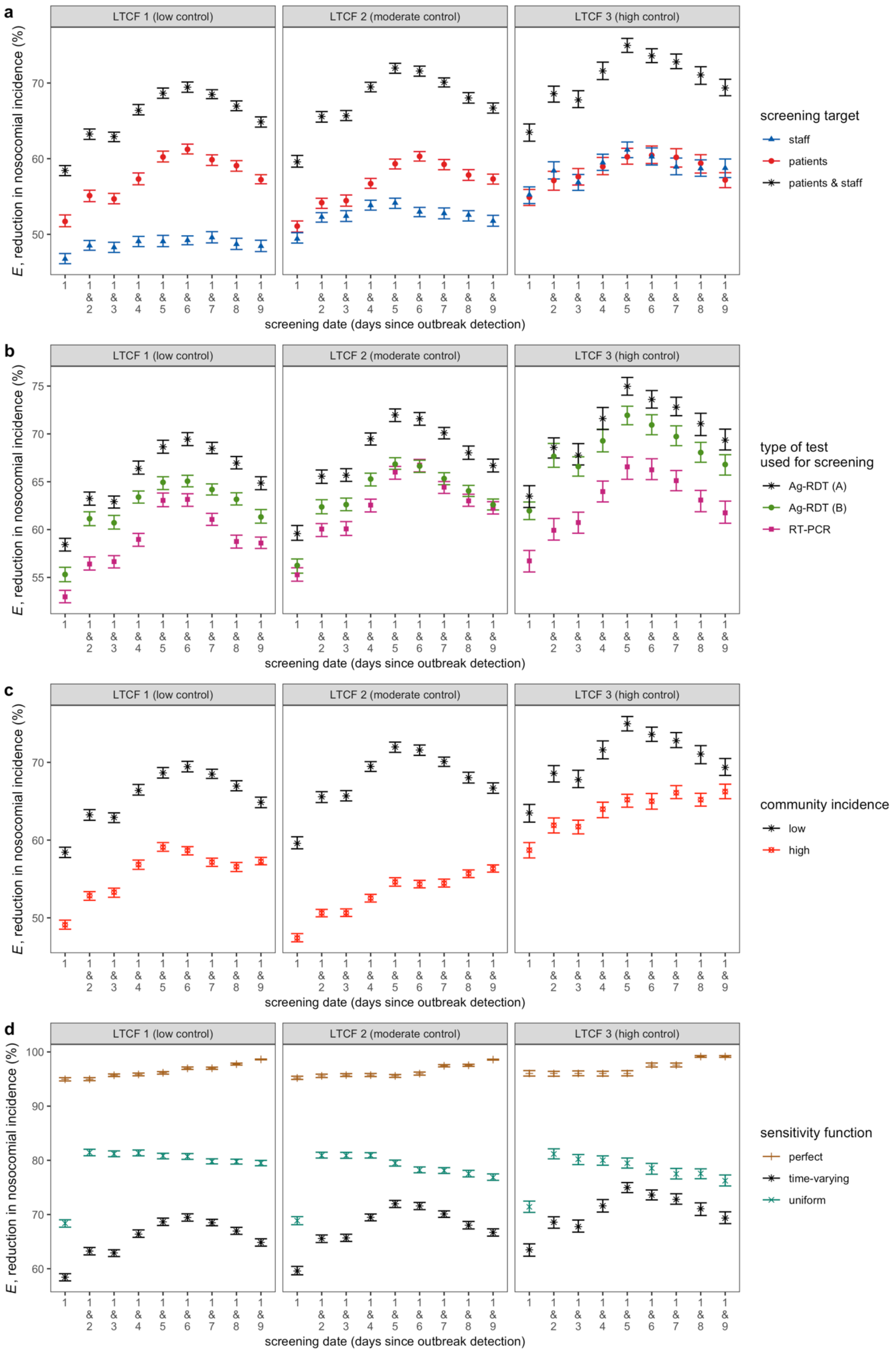


**Supplementary figure S9.** Sensitivity analyses: impacts of various model assumptions on efficacy of screening interventions. Points and error bars correspond to means and 95% confidence intervals estimated by bootstrap resampling (n=10,000). For all panels, black asterisks represent the assumption used in baseline analyses presented in the main text, unless specified otherwise. **(a)** Comparison of targeting patients and/or staff in screening interventions. **(b)** Comparison of using Ag-RDT or RT-PCR for screening, assuming immediate results for Ag-RDT and a 24-hour delay for RT-PCR. Two sensitivity curves for Ag-RDT are considered (see Supplementary figure S5). **(c)** Comparison of screening efficacy in the baseline low incidence scenario, and the alternative high incidence scenario. In LTCFs 2 and 3, optimal timing of second-round screening was delayed compared to baseline in the high incidence scenario. **(d)** Comparison of alternative sensitivity curves for both RT-PCR and Ag-RDT. When assuming uniform diagnostic sensitivity over the course of infection (70% for RT-PCR, 54% for Ag-RDT; turquoise points), the second round of screening was more effective the sooner it was conducted. Alternatively, assuming 100% diagnostic sensitivity for both RT-PCR and Ag-RDT (brown points), longer delays were more effective, with nosocomial incidence reduced by up to 98.6-99.2% with follow-up screening 8 days after the first round, the longest interval considered. Time-varying test sensitivity thus drives optimal efficacy of two-round screening at an intermediate screening lag. RT-PCR = reverse transcriptase polymerase chain reaction; Ag-RDT = antigen rapid diagnostic testing; LTCF = long-term care facility.

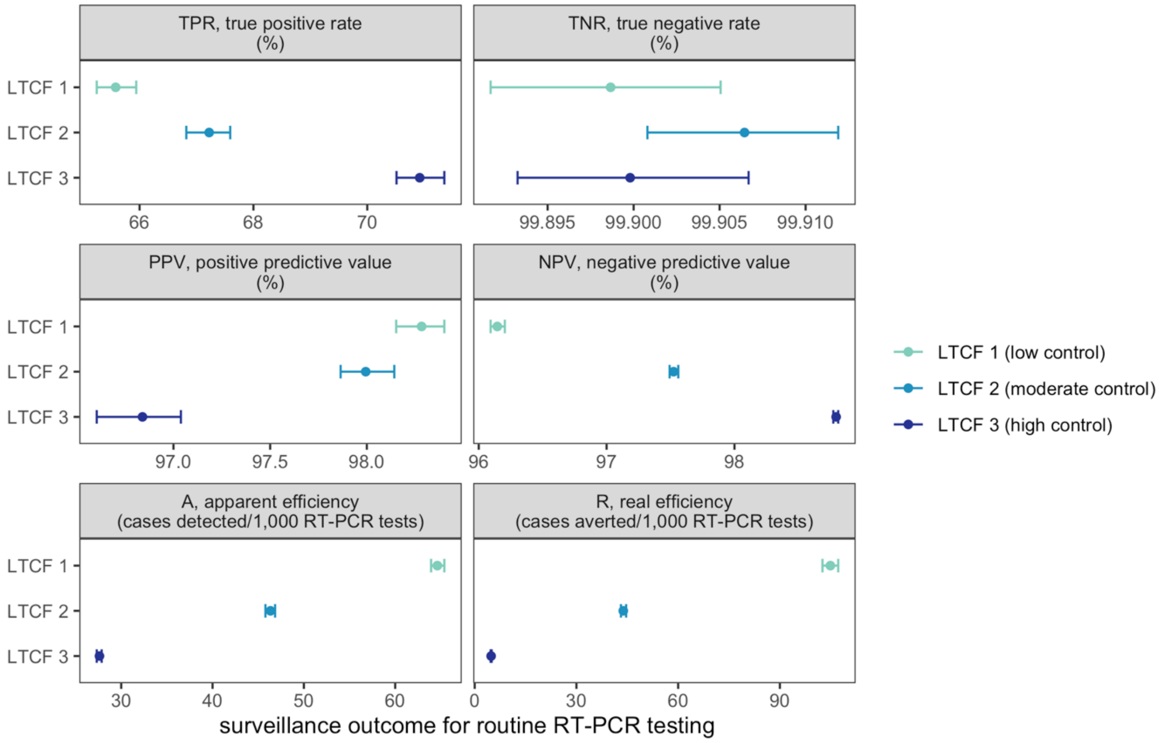


**Supplementary figure S10.** Performance of routine RT-PCR testing across LTCFs (y-axis, colours) and various surveillance outcomes (panels). Points and error bars correspond to means and 95% confidence intervals estimated by bootstrap resampling (n=10,000). RT-PCR = reverse transcriptase polymerase chain reaction; LTCF = long-term care facility.

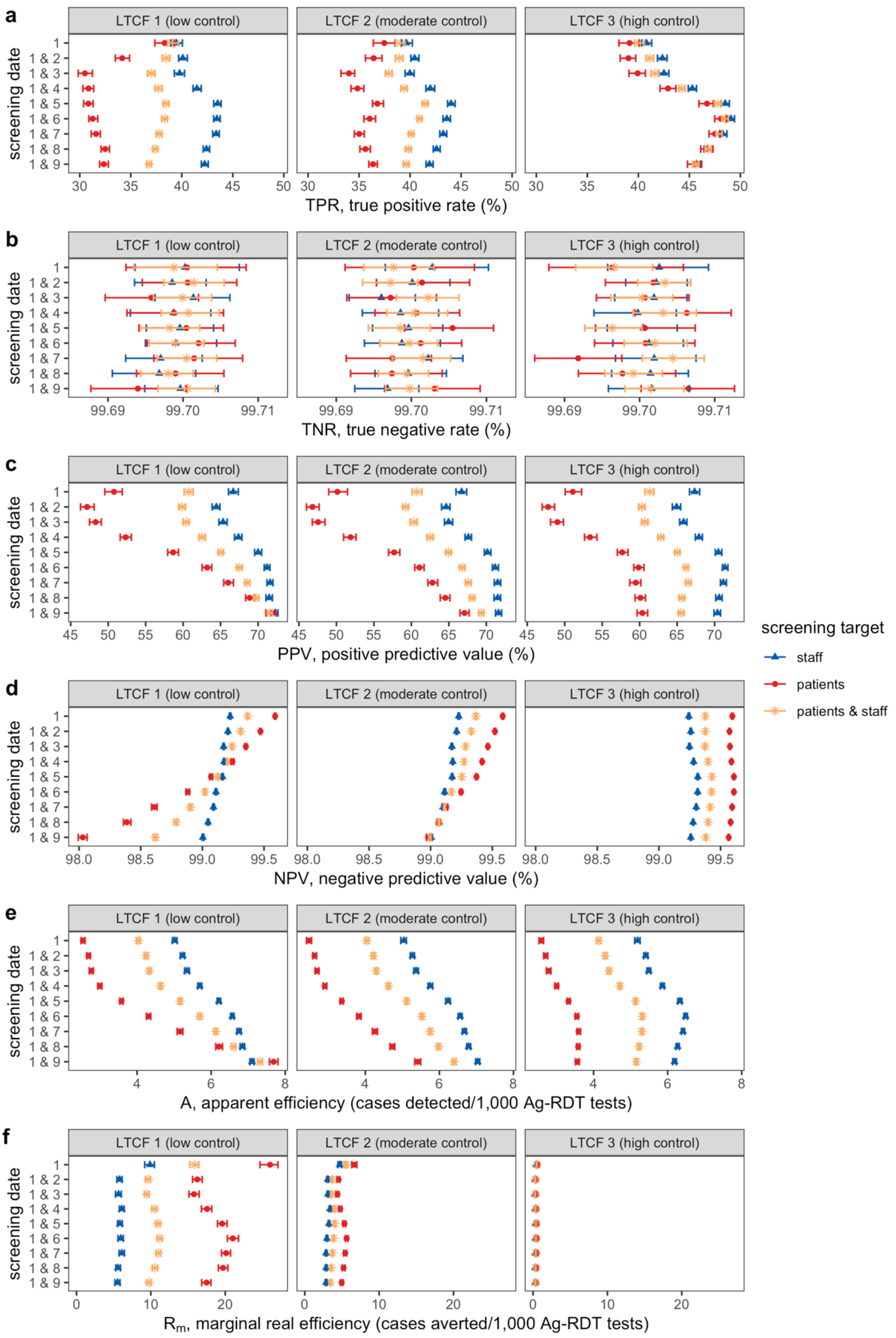


**Supplementary figure S11.** Performance of Ag-RDT screening interventions paired with routine RT-PCR testing (interventions #11 and #20 to #27 from Supplementary table S2) across various surveillance outcomes (rows). For **(f)**, note that efficacy reflects *marginal* cases averted: cases averted by Ag-RDT screening exclude cases already averted by routine RT-PCR testing. Points and error bars correspond to means and 95% confidence intervals estimated by bootstrap resampling (n=10,000). Ag-RDT = antigen rapid diagnostic testing; LTCF = long-term care facility.

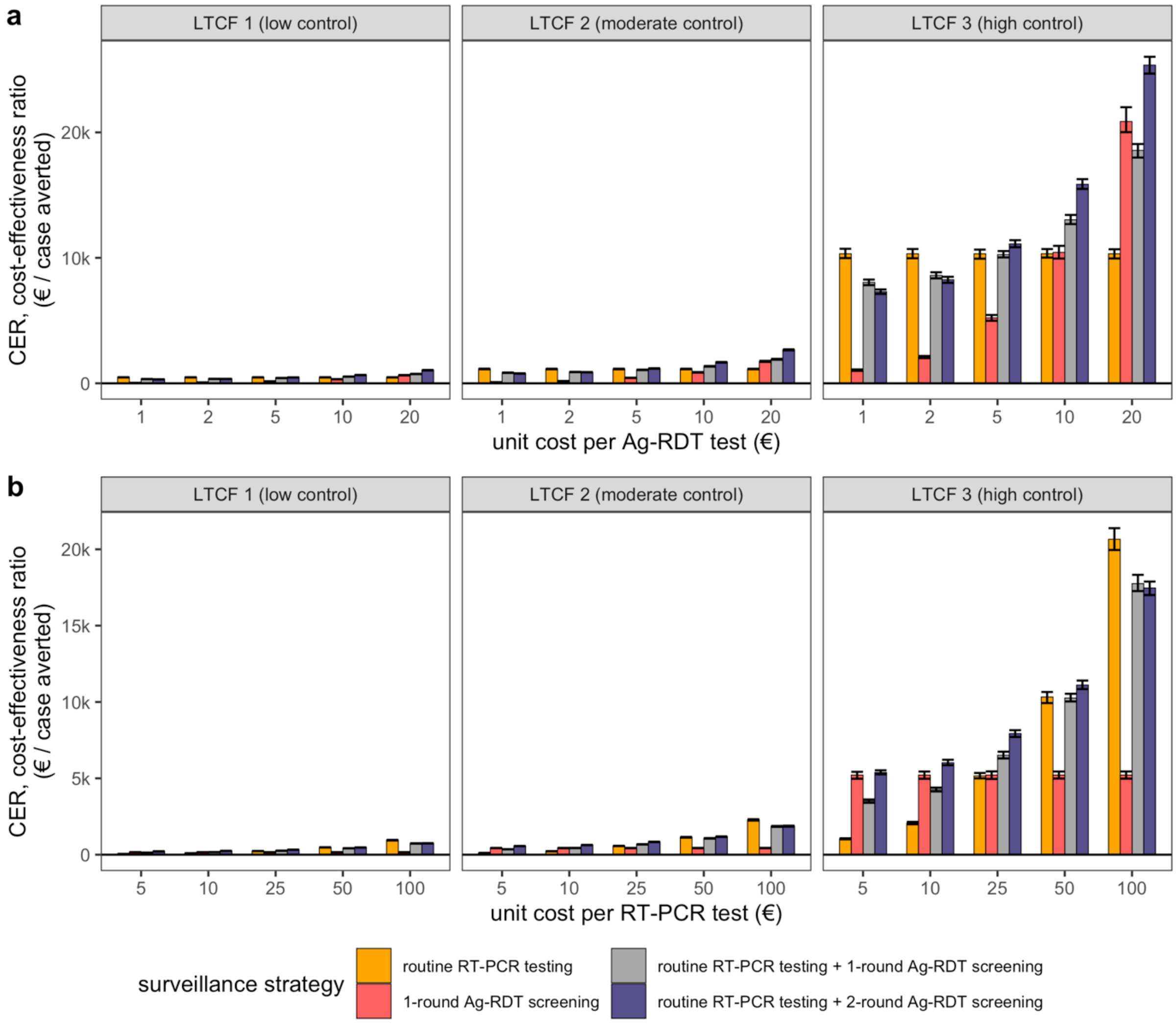


**Supplementary figure S12.** Cost-effectiveness ratios of four surveillance interventions (colours), estimated as surveillance unit costs per case averted. Cost-effectiveness was estimated while varying either **(a)** the unit cost per Ag-RDT test (at a fixed €50/RT-PCR test), or **(b)** the unit cost per RT-PCR test (at a fixed €5/Ag-RDT test). One-round screening was conducted on day 1 (interventions #2 and #11 in Supplementary table S2), and 2-round screening on days 1 and 5 (intervention #23). Baseline assumptions underlying simulations include: “low” community SARS-CoV-2 incidence; time-varying Ag-RDT sensitivity relative to RT-PCR (Ag-RDT A); and screening interventions that target all patients and staff in the LTCF. Bar heights and error bars correspond to means and 95% confidence intervals estimated by bootstrap resampling (n=10,000). RT-PCR = reverse transcriptase polymerase chain reaction; Ag-RDT = antigen rapid diagnostic testing; LTCF = long-term care facility.

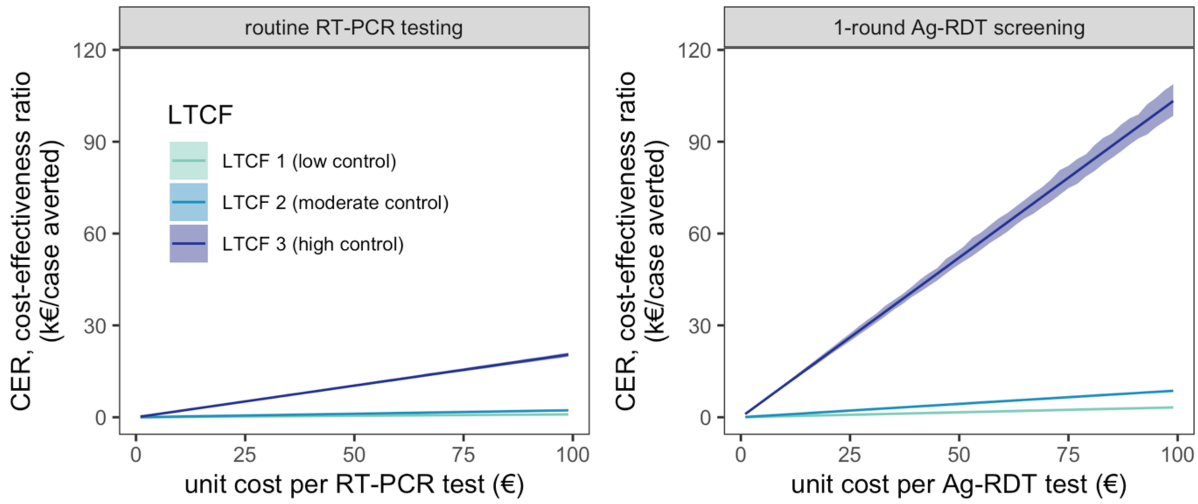


**Supplementary figure S13.** Cost-effectiveness ratios for routine RT-PCR (left, intervention #1 from Supplementary table S2) and 1-round Ag-RDT screening on day 1 (right, intervention #2 from Supplementary table S2) as a function of testing unit costs. Lines and shaded intervals correspond to means and 95% confidence intervals estimated by bootstrap resampling (n=10,000). RT-PCR = reverse transcriptase polymerase chain reaction; Ag-RDT = antigen rapid diagnostic testing; LTCF = long-term care facility.
